# Differences in transcriptional response underlie why prior vaccination exceeds prior infection in eliciting robust immune responses in Omicron infected outpatients

**DOI:** 10.1101/2022.03.24.22272837

**Authors:** Hye Kyung Lee, Ludwig Knabl, Mary Walter, Ludwig Knabl, Yuhai Dai, Magdalena Füßl, Yasemin Caf, Claudia Jeller, Philipp Knabl, Martina Obermoser, Christof Baurecht, Norbert Kaiser, August Zabernigg, Gernot M. Wurdinger, Priscilla A. Furth, Lothar Hennighausen

**Author notes:** **Correspondence:** HKL; LK; PAF; LH. These authors contributed equally to this work (Hye Kyung Lee, Ludwig Knabl, Priscilla A. Furth, Lothar Hennighausen). This author has senior authorship (Lothar Hennighausen).

## Abstract

Antibody response following Omicron infection is reported to be less robust than that to other variants. Here we investigated how prior vaccination and/or prior infection modulates that response. Disease severity, antibody responses and immune transcriptomes were characterized in four groups of Omicron-infected outpatients (n=83): unvaccinated/no prior infection, vaccinated/no prior infection, unvaccinated/prior infection and vaccinated/prior infection. The percentage of patients with asymptomatic or mild disease was highest in the vaccinated/no prior infection group (87%) and lowest in the unvaccinated/no prior infection group (47%). Significant anti-Omicron spike antibody levels and neutralizing activity were detected in the vaccinated group immediately after infection but were not present in the unvaccinated/no prior infection group. Within two weeks, antibody levels against Omicron, increased. Omicron neutralizing activity in the vaccinated group exceeded that of the prior infection group. No increase in neutralizing activity in the unvaccinated/no prior infection group was seen. The unvaccinated/prior infection group showed an intermediate response. We then investigated the early transcriptomic response following Omicron infection in these outpatient populations and compared it to that found in unvaccinated hospitalized patients with Alpha infection. Omicron infected patients showed a gradient of transcriptional response dependent upon prior vaccination and infection status that correlated with disease severity. Vaccinated patients showed a significantly blunted interferon response as compared to both unvaccinated Omicron infected outpatients and unvaccinated Alpha infected hospitalized patients typified by the response of specific gene classes such as OAS and IFIT that control anti-viral responses and IFI27, a predictor of disease outcome.

## Introduction

The highly transmissible Omicron (B.1.1.529) variant is less susceptible to neutralizing antibodies elicited by previous vaccination or other variant infection (1-4), thus resulting in a continuation of the COVID-19 pandemic. While recent studies have investigated the antibody response to breakthrough infections (5, 6), there is a knowledge gap about the humoral and genomic immune response to Omicron infection in outpatients that had been vaccinated, previously infected with another SARS-CoV-2 variant or both. Specifically, the impact of previous infection as compared to vaccination in the generation of anti-Omicron spike antibodies upon Omicron infection has yet to be determined.

While Omicron infections generally result in a more moderate symptomology compared to other variants, the genomic immune response in this patient population has not been investigated. Transcriptome studies on hospitalized patients infected with the Alpha (7) or Beta (8) variants have revealed the activation of interferon pathway genes with an emphasis of the JAK/STAT pathway. While the interferon response in severely ill patients has been reported, it is not clear if Omicron patients with a generally lighter symptomology have a different immune transcriptome which can be further modulated by vaccination and prior infection.

To address these questions, we investigate the transcriptional and humoral immune response in 83 outpatients with documented Omicron infection. Thirty-four had no prior infection and were not vaccinated, 23 persons had been vaccinated and boosted with the BNT162b2 mRNA vaccine (Pfizer–BioNTech), 19 had a history of prior infection by other SARS-CoV-2 variants and 7 had been vaccinated and had a prior infection. This study permitted an understanding of how vaccination and prior infection impact the immune response to Omicron infection and we identified a gradient response that corresponds to the humoral profile.

## Results

### Study design

The first case of Omicron infection in Austria was reported at the end of November 2021 (9) and recruitment of outpatients took place between December 2021 and March 2022. Here, we investigate the humoral and transcriptional immune response in 83 persons with documented Omicron infection. Four groups were studied: no vaccination/no prior infection (n=34), vaccination/no prior infection (n=23), no vaccination/prior infection (n=19) and vaccination/prior infection (n=7) (Figure 1A; Table 1). Demographic and clinical characteristics of the study population are provided in Table 1. The transcriptional response of the first three groups was investigated at days 1-3 and the humoral response at two timepoints (days 1-3 and 12-15). The humoral response for the fourth group (vaccination/prior infection) was measured at days 12-15 for comparison. Due to recruitment challenges and to the low numbers of these people in the community studied, only samples for the later timepoint were available for that group. Transcriptional studies were limited to days 1-3, the point of highest immune transcriptional response. The percentage of patients with asymptomatic or mild disease without respiratory symptoms was highest in the vaccination/no prior infection group (87%), followed by the no vaccination/prior infection group (63% with mild disease, no asymptomatic), the no vaccination/no prior infection (47% with either mild disease or asymptomatic) group and the vaccination/prior infection (72% with either mild disease or asymptomatic) group (Figure 1B). Moderate and severe cases accounted for 24% in the no vaccination/no prior infection group but only 4% of the vaccinated group, 11% in the no vaccination/prior infection group and 14% in the vaccinated/prior infection group (Figure 1B).

**Figure 1.**
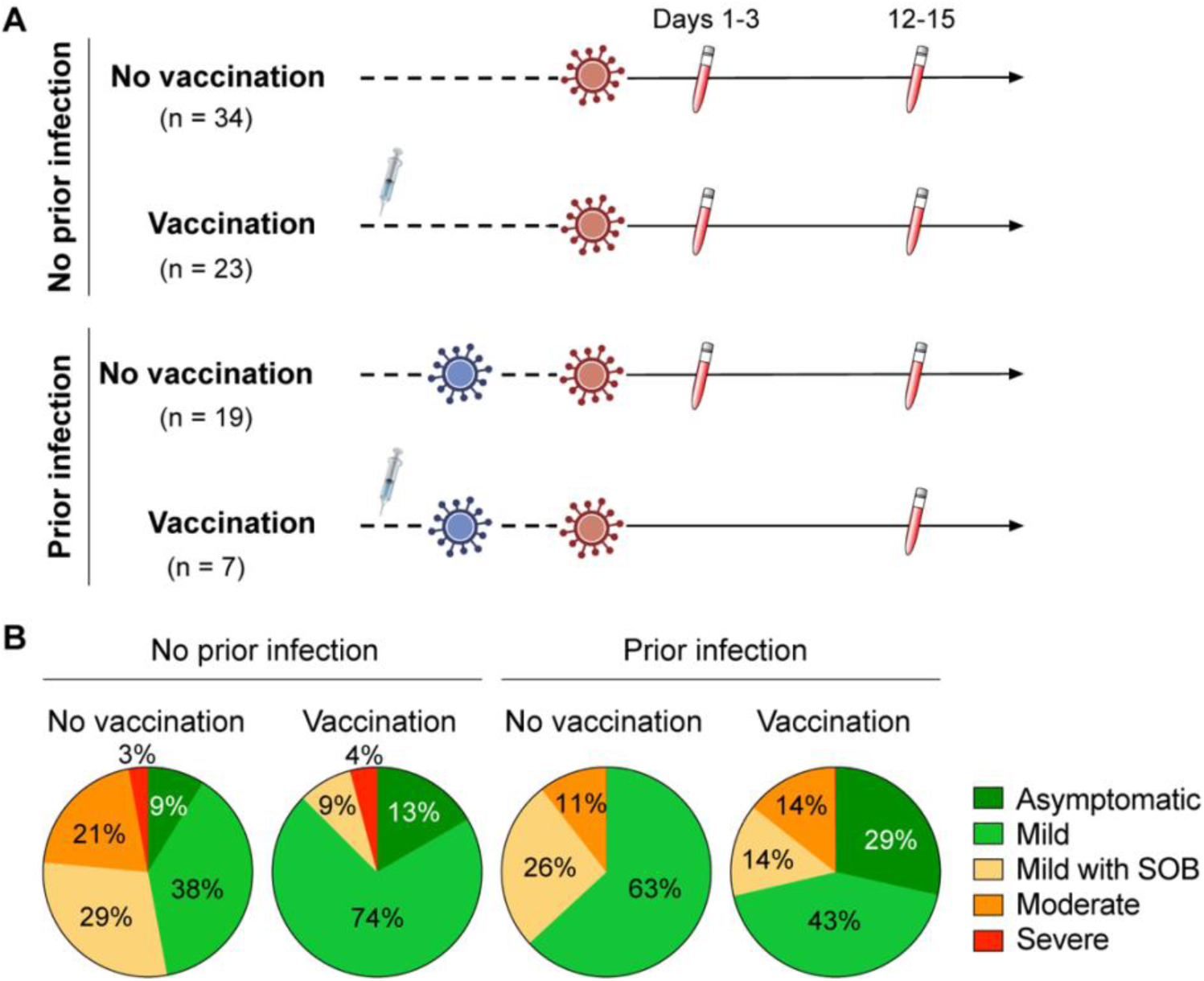
Study design and outpatient symptomology. **(A)** Schematic presentation of the experimental workflow. All 83 study subjects were infected by the SARS-CoV-2 Omicron variant with or without prior infection with another SARS-CoV-2 variant. Four groups were studied: no vaccination/no prior infection (n=34), vaccination/no prior infection (n=23), no vaccination/prior infection (n=19) and vaccination/prior infection (n=7) (Table 1). Blood was collected from the outpatients from three groups at two timepoints and from the fourth group at one time point after testing PCR positive. **(B)** Symptomology of the study cohorts (Table 1). SOB, shortness of breath.

**Table 1.**
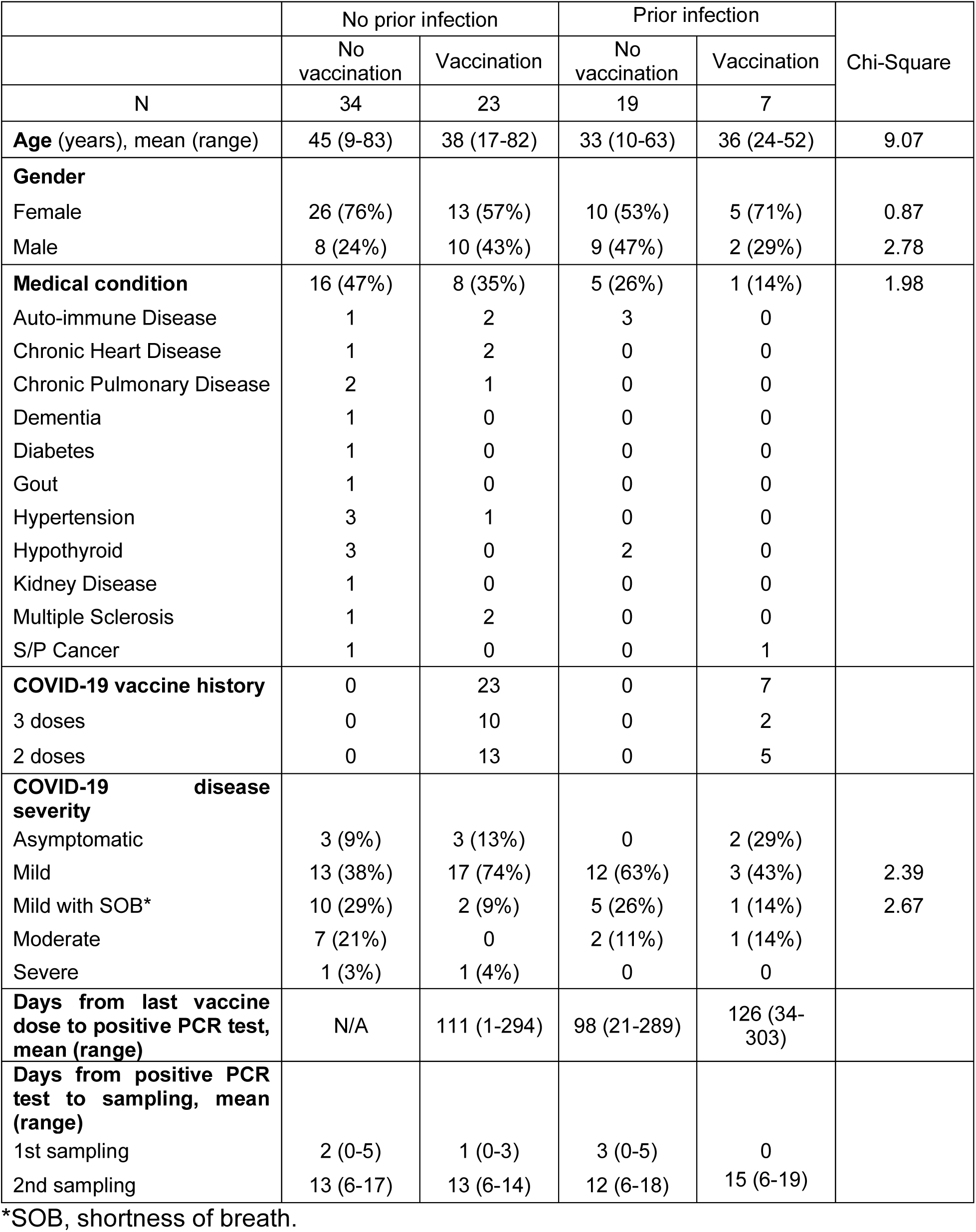
Characteristics of Omicron study population.

### Vaccination is superior to prior infection in preparing the immune response to Omicron

First, we measured circulating anti-spike antibody levels in serum samples obtained from the Omicron outpatients within the first three days (Day 1-3) of validated infection (Figure 2). The anti-Omicron spike IgG levels were highest in the vaccinated patients without prior infection and approximately 10-fold lower in the unvaccinated patients with and without prior infection (Figure 2A). An equivalent pattern was obtained for the ancestral strain (Figure 2A) and other variants (Figure S1A). A significant increase of anti-Omicron spike, but not anti-ancestral spike, antibodies was observed in the vaccinated and the previously infected group within 12-15 days following Omicron infection (Figure 2A). Antibody levels did not increase in the naïve group. These findings are mirrored by anti-spike antibody levels from other variants (Figure S1A).

**Figure 2.**
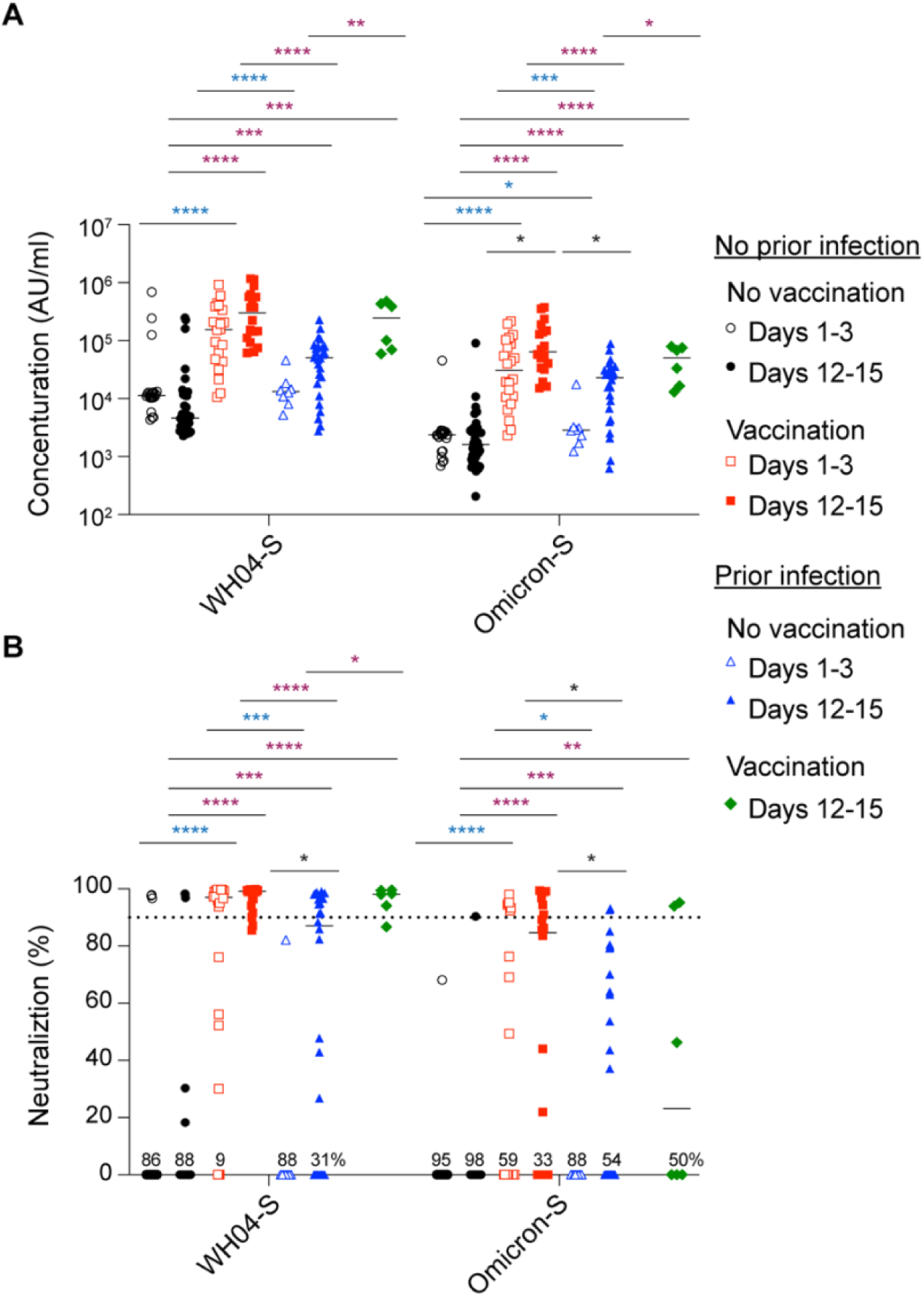
Antibody analysis. **(A)** Plasma IgG antibody binding the SARS-CoV-2 RBD (spike) from the ancestral and Omicron strains in the unvaccinated and vaccinated Omicron patients without or with prior infection experience. **(B)** Neutralizing antibody response to virus spike protein of the ancestral and Omicron variants. *p*-value between two groups is from one-tailed Mann-Whitney t-test. Black Asterix shows significance between days in the same group, blue ones do significance of days 1-3 between groups, and purple ones do significance of days 12-15 between groups. \**p* < 0.05, \*\**p* < 0.01, \*\*\**p* < 0, \*\*\*\**P* < 0.0001. Line at median.

At this point in the pandemic, a critical question is whether prior BNT162b2 vaccination can prompt development of neutralizing antibodies in Omicron infected individuals. Here, we assessed neutralization capacity using the angiotensin-converting enzyme 2 (ACE2) binding inhibition assay, against the Omicron spike protein and those from other variants. We measured neutralization within three days after validated Omicron infection (Days 1-3) and after 12-15 days (Figure 2B). Significant (approximately 11-fold higher in the vaccinated group compared to the no vaccinated group) Omicron neutralizing activity was seen in the vaccinated group, which further increased after 12-15 days. Significant neutralizing activity was detected in the previously infected group, with diverse increase, but less than the vaccinated group, at day 12-15. These findings parallel those seen for other variants (Figure S1B).

### Vaccination but not prior infection blunts interferon responses elicited by Omicron infection

To understand the impact of prior vaccination or prior infection on the genomic immune response to Omicron infection, we investigated the immune transcriptome (Figure 3; Table S1). These data sets were also compared to a naïve reference population (no previous SARS-CoV-2 infection and no vaccination) (10) and to hospitalized patients that had been infected with the Alpha variant (7). Bulk RNA-seq on buffy coats isolated within the first two days after validated Omicron infection was performed with an average sequencing depth of 200 million reads per sample. First, we directly compared the transcriptomes of the vaccinated and unvaccinated Omicron cohorts without prior infection with the control cohort of 30 healthy individuals from the same geographic area (Tyrol Control Transcriptomes, TCT). Expression of 489 and 732 genes was induced significantly in the no vaccination and vaccination group, respectively. Expression of 146 and 246 genes was reduced. Next, we compared the transcriptomes between TCT and no vaccination/prior infection group and found 356 significantly induced and 153 reduced genes. GSEA analyses linked the induced genes in three groups to innate immune responses including interferon response and cytokine signaling through the JAK/STAT pathway (Figure 3A). An even stronger enrichment of innate immune genes was observed in hospitalized patients infected with Alpha variant (Figure 3A). Overall, the differences observed between the cohorts were of a quantitative rather than a qualitative nature.

**Figure 3.**
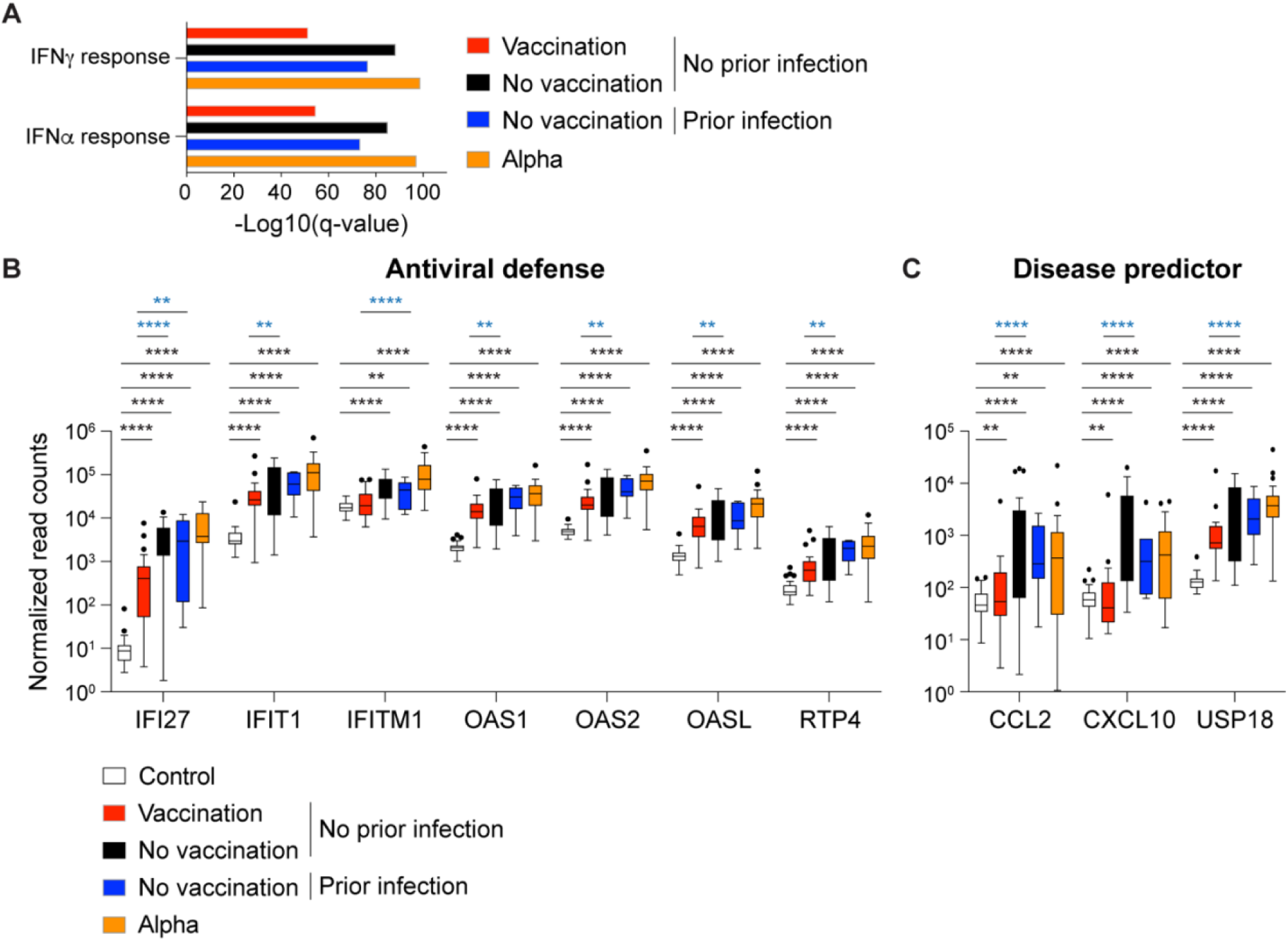
Immune transcriptomes following Omicron infection. **(A)** Gene categories expressed at significantly higher levels in unvaccinated and vaccinated Omicron outpatients with or without prior SARS-CoV-2 infection, and in COVID-19 patients infected by the Alpha variant were significantly enriched in interferon-activated and inflammatory pathways. X-axis denotes statistical significance as measured by minus logarithm of FDR q-values. Y-axis ranked the terms by q values. **(B-C)** Bar plots with the relative mRNA levels of fifteen innate immune response genes of the 23 genes that are significantly induced in all cohorts and higher in the no vaccination group compared to vaccination group (B) and seven that are significantly induced in No vaccination group, but not Vaccination group, and higher in No vaccination group compared to Vaccination group (C). Black Asterix shows significance between healthy control and COVID-19 groups and blue ones do significance between Omicron groups.

We directly explored those genes that were differentially expressed between vaccinated and unvaccinated outpatients depending on their prior infection status. (Figure 3B, Table S1). In general, the activation of genes linked to antiviral defense programs and COVID-19 disease predictors was lowest in the vaccinated patients without prior infection and highest in the patients infected with the Alpha variant (Figure 3B-C). The antiviral defense programs encompass interferon-induced gene families, including the OAS family that counteract viral attacks by degrading viral RNAs. We also identified genes that had not previously linked to COVID-19, such as USP18, an IFN-induced gene encoding a negative regulator of type I IFN signaling (11), which highly activated in unvaccinated Omicron patients and Alpha patients.

Of particular interest is the OAS family of antiviral enzymes where hypomorphic mutations have been associated with susceptibility to viral infection and activating mutations with autoimmune disease. The OAS1 member of this family is of particular interest as it harbors a mutation traced back to the Neanderthal genome that results in a splice variant associated with protection from severe COVID-19 (12). RNA-seq data revealed that the rs10774671 haplotype was found in 83% of our control population (TCT cohort), 86% and 85% in the unvaccinated and vaccinated Omicron population, respectively and 96% of the Alpha population. Induction of OAS1 expression is highest in unvaccinated Omicron patients and the Alpha patients (Figure 3B).

## Discussion

Here, we demonstrate that prior vaccination with BNT162b2 modifies the transcriptional immune response to Omicron breakthrough infections and induces a more robust Omicron antibody response. Specifically, even among non-hospitalized patients there is gradient response dependent on previous immunological status.

Omicron infection of unvaccinated and vaccinated people has been reported to result in milder disease relative to previous variants (13). However, unlike the response to Delta breakthrough infection (5, 6, 14), Omicron breakthrough infections might result in lower levels of neutralizing antibodies (15). The muted antibody response may be due to the high share of asymptomatic and mild infections as is also indicated by a less active immune transcriptome shown in our study. Notably, individuals with prior vaccination showed significant Omicron neutralization activity, even in the presence of a blunted transcriptional response. Naïve individuals demonstrated significantly higher transcriptional response but a less robust humoral response. The response in vaccinated or prior infection groups was quantitatively less, but similar in unvaccinated/no prior infection group, when compared to the transcriptional response of hospitalized individuals with Alpha infection.

Vaccination was vastly superior to prior infection in promoting neutralizing Omicron antibodies and subsequent Omicron infection yielded an elevated antibody response in both groups. In contrast, Omicron infection in the antigen naïve population did not result in any significant antibody production within a time window of two weeks, suggesting that the initial exposure to Omicron spike proteins does not elicit a substantial immune response.

Hospitalized COVID-19 patients show powerful transcriptomic responses in peripheral blood with conserved genetic components that are comprehensively interferon-driven. A qualitatively similar, yet quantitative distinct, transcriptional response was observed in outpatients infected with Omicron. The blunted activity of interferon response pathways, especially in the vaccinated group, mirrors the overall mild symptomology seen in our patients. As expected, expression of COVID-19 disease predictors was greatly reduced compared to the hospitalized patients infected with Alpha.

In summary, vaccination prior to Omicron infection modifies humoral and transcriptional responses with higher antibody and neutralization titers and lower interferon gamma activation than that found in unvaccinated individuals either with or without prior infection.

## Methods

### Ethics

This study was approved (EK Nr: 1064/2021) by the Institutional Review Board (IRB) of the Office of Research Oversight/Regulatory Affairs, Medical University of Innsbruck, Austria, which is responsible for all human research studies conducted in the State of Tyrol (Austria). Participant information was coded and anonymized.

### Study population, study design and recruitment

A total of 83 patients infected with Omicron were recruited for the study under informed consent. Thirty-four with no history of prior vaccination or prior infection with other SARS-CoV-2 variants, 23 patients who had received 2 or 3 doses of the BNT162b2 vaccine, 19 unvaccinated patients with prior infections and 7 vaccinated patients with prior infection (Table 1). Recruitment and blood sample collection took place between December 2021 and March 2022. This study was approved (EK Nr: 1064/2021) by the Institutional Review Board (IRB) of the Office of Research Oversight/Regulatory Affairs, Medical University of Innsbruck, Austria, which is responsible for all human research studies conducted in the State of Tyrol (Austria). The investigators do not need to have an affiliation with the University of Innsbruck. A waiver of informed consent was obtained from the Institutional Review Board (IRB) of the Office of Research Oversight/Regulatory Affairs, Medical University of Innsbruck (https://www.i-med.ac.at/ethikkommission/). **Written informed consent was obtained from all subjects**. This study was determined to impose minimal risk on participants. All methods were carried out in accordance with relevant guidelines and regulations. All research has been have been performed in accordance with the Declaration of Helsinki (https://www.wma.net/policies-post/wma-declaration-of-helsinki-ethical-principles-for-medical-research-involving-human-subjects/). In addition, we followed the [‘Sex and Gender Equity in Research – SAGER – guidelines’](https://researchintegrityjournal.biomedcentral.com/articles/10.1186/s41073-016-0007-6) and included sex and gender considerations where relevant.

### Antibody assay

End-point binding IgG antibody titers to various SARS-CoV-2–derived antigens were measured using the Meso Scale Discovery (MSD) platform. SARS-CoV-2 spike, nucleocapsid, Alpha, Beta, Gamma, Delta, and Omicron spike subdomains were assayed using the V-plex multispot COVID-19 serology kits (Panel 23 (IgG) Kit, K15567U). Plates were coated with the specific antigen on spots in the 96 well plate and the bound antibodies in the samples (1:50000 dilution) were then detected by anti-human IgG antibodies conjugated with the MSD SULPHO-TAG which is then read on the MSD instrument which measures the light emitted from the tag.

### ACE2 binding inhibition (Neutralization) ELISA

The V-PLEX COVID-19 ACE2 Neutralization kit (Meso Scale Discovery, Panel 23 (ACE2) Kit, K15570U) was used to quantitatively measure antibodies that block the binding of ACE2 to its cognate ligands (SARS-CoV-2 and variant spike subdomains). Plates were coated with the specific antigen on spots in the 96 well plate and the bound antibodies in the samples (1:10 dilution) were then detected by Human ACE2 protein conjugated with the MSD SULPHO-TAG which is then read on the MSD instrument which measures the light emitted from the tag.

### Extraction of the buffy coat and purification of RNA

Whole blood was collected, and total RNA was extracted from the buffy coat and purified using the Maxwell RSC simply RNA Blood Kit (Promega) according to the manufacturer’s instructions. The concentration and quality of RNA were assessed by an Agilent Bioanalyzer 2100 (Agilent Technologies, CA).

### mRNA sequencing (mRNA-seq) and data analysis

The Poly-A containing mRNA was purified by poly-T oligo hybridization from 1 mg of total RNA and cDNA was synthesized using SuperScript III (Invitrogen, MA). Libraries for sequencing were prepared according to the manufacturer’s instructions with TruSeq Stranded mRNA Library Prep Kit (Illumina, CA, RS-20020595) and paired-end sequencing was done with a NovaSeq 6000 instrument (Illumina) yielding 200-350 million reads per sample.

The raw data were subjected to QC analyses using the FastQC tool (version 0.11.9) (https://www.bioinformatics.babraham.ac.uk/projects/fastqc/). mRNA-seq read quality control was done using Trimmomatic (16) (version 0.36) and STAR RNA-seq(17) (version STAR 2.5.4a) using 150 bp paired-end mode was used to align the reads (hg19). HTSeq (18) (version 0.9.1) was to retrieve the raw counts and subsequently, Bioconductor package DESeq2 (19) in R (https://www.R-project.org/) was used to normalize the counts across samples and perform differential expression gene analysis. Additionally, the RUVSeq(20) package was applied to remove confounding factors. The data were pre-filtered keeping only genes with at least ten reads in total. The visualization was done using dplyr (https://CRAN.R-project.org/package=dplyr) and ggplot2 (21). The genes with log2 fold change >1 or <-1 and adjusted p-value (pAdj) <0.05 corrected for multiple testing using the Benjamini-Hochberg method were considered significant and then conducted gene enrichment analysis (GSEA, https://www.gsea-msigdb.org/gsea/msigdb).

### Quantification and statistical analysis

Differential expression gene (DEG) identification used Bioconductor package DESeq2 in R. P-values were calculated using a paired, two-side Wilcoxon test and adjusted p-value (pAdj) corrected using the Benjamini–Hochberg method. Genes with log_2_ fold change >1 or <-1, pAdj <0.05 and without 0 value from all sample were considered significant. For significance of each GSEA category, significantly regulated gene sets were evaluated with the Kolmogorov-Smirnov statistic. P-values of antibody between two groups were calculated using one-tailed Mann-Whitney t-test on GraphPad Prism software (version 9.0.0). A value of \**P* < 0.05, \*\**P* < 0.01, \*\*\**P* < 0.001, \*\*\*\**P* < 0.0001 was considered statistically significant.

## Data Availability

RNA-seq data from buffy coat of healthy control and COVID-19 Alpha patients were obtained GSE189039, GSE190747 and GSE190680. The RNA-seq data from this study will be uploaded in GEO before publishing the manuscript.

## Author Contributions

HKL, LK, PAF and LH designed the study. LK Sr., MF, YC, CJ, PK, MO, CB, NK, AZ and GMW recruited patients and collected material. HKL analyzed RNA-seq data. MW and YD conducted IgG antibody and neutralization assay. HKL, PAF and LH analyzed data. HKL administrated project. HKL, PAF, and LH wrote the paper. All authors read and approved the manuscript.

## Acknowledgments

Our gratitude goes to the participants who contributed to this study to advance our understanding of Omicron and COVID-19. This work was utilized the computational resources of the NIH HPC Biowulf cluster (http://hpc.nih.gov). RNA-sequencing was conducted in the NIH Intramural Sequencing Center, NISC (https://www.nisc.nih.gov/contact.htm).

